# Multi-dimensional attention framework for personalized Alzheimer’s disease progression prediction across sporadic and genetic risk cohorts

**DOI:** 10.64898/2025.12.13.25342199

**Authors:** Zhiyuan Song, Sarah E. Morgan, Shahid Zaman

## Abstract

Alzheimer’s disease manifests through heterogeneous progression patterns across diverse populations, yet current predictive models fail to capture this complexity while maintaining clinical interpretability. Here we present a multi-dimensional attention framework that simultaneously captures temporal dynamics and biomarker importance patterns to predict disease progression across three fundamentally different populations: the general late-onset population using the Alzheimer’s Disease Prediction Of Longitudinal Evolution (TADPOLE) dataset (N=1669), cases with Down Syndrome-associated Alzheimer’s disease using the Alzheimer’s Biomarker Consortium - Down Syndrome (ABC-DS) dataset (N=396) and cases with autosomal dominant Alzheimer’s disease using the Dominantly Inherited Alzheimer Network (DIAN) dataset (N=425). Our framework achieved multi-class area under the ROC curve (mAUC) values of 0.793 (TADPOLE), 0.680 (ABC-DS) and 0.902 (DIAN) when trained on each dataset independently to predict individuals’ future diagnostic status (cognitively normal, mild cognitive impairment, or Alzheimer’s disease) based on their longitudinal biomarker history, outperforming conventional approaches. The model generates individual-specific attention maps revealing distinct biomarker importance over time. Transfer learning from the TADPOLE dataset-which included neuroimaging data-improved prediction performance on the ABC-DS dataset with no neuroimaging data included from 0.680 to 0.771, demonstrating that disease mechanisms transcend both etiological boundaries and data modalities. Ultimately this framework could help enable precision medicine approaches for data-limited cohorts across the Alzheimer’s disease spectrum.

## Introduction

Alzheimer’s disease (AD) is a neurodegenerative disorder affecting over 55 million people worldwide [1]. While the general population typically develops late-onset disease after age 65, genetically at-risk groups experience illness much earlier: individuals with Down syndrome show pathology by age 40, while autosomal dominant mutation carriers develop symptoms in their 40s to 50s [2] [3]. However, disease-modifying treatments remain limited across all populations [4]. The disease’s heterogeneity presents fundamental challenges for prognosis and treatment, emphasising the need for personalized strategies. Accurate early prediction across the diagnostic spectrum (cognitively normal (CN), mild cognitive impairment (MCI) and Alzheimer’s disease (AD)) could support timely, targeted intervention [4].

The Alzheimer’s Disease Neuroimaging Initiative (ADNI) [5] and similar modern cohorts for sporadic AD provide unprecedented multimodal longitudinal data encompassing neuroimaging, cerebrospinal fluid biomarkers, genetic markers and cognitive assessments to develop models to predict disease progression in the general population. However, genetically at-risk populations, including those with Down syndrome and autosomal dominant mutations, remain understudied with smaller datasets and limited multimodal data. This disparity in data availability raises important questions about whether predictive approaches developed for well-characterized sporadic cohorts can be adapted to improve outcomes in data-limited genetic populations. The high-dimensional nature of these datasets challenges traditional models, requiring advanced computational approaches with clinical interpretability.

Machine learning methods have shown promise for Alzheimer’s disease research, achieving high accuracy in cross-sectional diagnostic classification, although longitudinal disease progression prediction remains challenging [6] [7] [8] [9]. For example, studies classifying AD versus controls using vision transformers achieved pooled sensitivities (the average true positive rate across multiple test sets) exceeding 92% [9] [10] [11]. However, predicting longitudinal progression is substantially more difficult due to heterogeneous progression trajectories, where disease progression rates vary substantially between individuals, with some patients declining rapidly while others remain stable for years [6] [12]. Additionally, biomarker trajectories follow nonlinear patterns where the predictive value of specific markers changes across disease stages [6]. While prior approaches using traditional machine learning and recurrent neural networks have been applied to longitudinal AD data [8], they lack mechanisms to capture how biomarker importance evolves over time and to provide individual-level interpretability. Time-Aware Long Short-Term Memory networks (LSTMs) incorporate elapsed time between assessments and show high potential for other clinical prediction tasks [13] [14]. For example, the RETAIN model pioneered dual attention identifying influential timepoints and variables, achieving an AUC of 0.8705 for heart failure prediction [15].

Another key challenge is that most approaches have not been validated across populations with different disease etiologies, limiting understanding of whether predictive patterns generalize across genetic backgrounds [6] [7]. Different AD populations share core pathophysiology despite distinct etiologies [16] [17]. While TADPOLE captures late-onset sporadic disease [18], genetically defined populations follow distinct temporal patterns: Down syndrome individuals develop early-onset AD from chromosome 21 triplication [19], and autosomal dominant AD from amyloid precursor protein (APP), presenilin 1 (PSEN1), or presenilin 2 (PSEN2) mutations follows predictable progression [20]. Despite different onset ages, the pathological sequence (amyloid accumulation, tau propagation, neurodegeneration) remains consistent [16]. This conservation suggests that transfer learning, where models trained on large well-characterized datasets are adapted to improve predictions in smaller cohorts [21], has high potential across AD populations, scientifically revealing shared disease mechanisms while enabling model application to data-limited populations. This approach particularly benefits genetically at-risk populations where comprehensive data collection faces practical barriers, for example individuals with Down syndrome often experience anxiety during neuroimaging procedures, making extensive scanning protocols challenging [19] [22]. Meanwhile, autosomal dominant AD cohorts remain inherently small due to mutation rarity, resulting in limited sample sizes for model training [20] [23].

Finally, achieving clinical utility may require not only accurate predictions but also interpretability at the individual patient level, particularly regarding how feature importance evolves over disease progression [24]. Attention mechanisms offer a promising approach to this challenge by quantifying the relative contribution of different biomarkers at each disease stage [25]. Understanding which features drive predictions for individual patients and how their importance changes over time remains a critical gap. Prior work has shown that feature importance varies substantially across disease stages [7], yet most existing deep learning models for AD progression typically capture these temporal dynamics of biomarker relevance only at the population level, without providing individual-specific insights. Structural imaging features may dominate early predictions while cognitive measures become increasingly important in later stages [26]. This temporal evolution of feature importance has direct implications for personalized monitoring strategies and treatment decisions. Our framework helps address this need by generating individual-specific attention maps that reveal biomarker importance over time.

Our study addresses three key questions: (1) Can a unified attention architecture predict longitudinal AD outcomes across general and genetic populations with high accuracy? (2) Can transfer learning from general populations improve prediction accuracy for data-limited genetic cohorts? (3) Which features drive predictions for different populations, how do they change over time, and are there common themes across populations? Ultimately, we present a multidimensional attention framework combining temporal and feature attention to predict AD progression across TADPOLE (sporadic), ABC-DS (Down syndrome), and DIAN (autosomal dominant). The framework provides interpretable insights while achieving state-of-the-art prediction accuracy across genetically diverse populations.

## Methods

### 1.1 Datasets

We utilized three independent cohorts to train and evaluate our multi-dimensional attention pipeline, each representing a distinct population for Alzheimer’s disease. Details are given in **Table 1** and below:

**Table 1:** Demographic and clinical characteristics of the three study cohorts. The table presents data from the Alzheimer’s Disease Prediction of Longitudinal Evolution (TADPOLE) dataset representing late-onset sporadic Alzheimer’s disease, the Alzheimer’s Biomarker Consortium - Down Syndrome (ABC-DS) dataset representing Down syndrome-associated Alzheimer’s disease, and the Dominantly Inherited Alzheimer Network (DIAN) dataset representing autosomal dominant Alzheimer’s disease. For each dataset, participants are stratified by diagnostic group: cognitively normal controls, mild cognitive impairment (MCI), and Alzheimer’s disease (AD). Available data modalities for each cohort are shown, including magnetic resonance imaging (MRI) regions of interest (ROIs) with volumetric and cortical measures, positron emission tomography (PET) imaging with fluorodeoxyglucose (FDG), florbetapir (AV45), and flortaucipir (AV1451) tracers, diffusion tensor imaging (DTI) ROIs, cerebrospinal fluid (CSF) biomarkers, apolipoprotein E (APOE) genotyping, karyotyping for chromosomal analysis, and various cognitive and neurological assessments.

| **Dataset** |  |  |  |  |  | **Modalities** |
| --- | --- | --- | --- | --- | --- | --- |
| **TADPOLE,** | Subject group | Number (% of all subjects) | Visits per subject | Age | Gender (%female) | Demographics<br>Diagnosis<br>Cognitive tests |
|  | Controls | 502 (30.1%) | 7.0±4.1 | 73.6±6.2 | 52.0% | MRI ROIs (volumes, cortical thicknesses, |
| **total N =**<br>**1669** | MCI | 617 (37.0%) | 7.0±3.7 | 73.2±7.6 | 40.2% | surface areas)<br>FDG PET<br>AV45 PET<br>AV1451 PET<br>DTI ROIs<br>CSF<br>APOE genotype |
|  | AD | 550 (33.0%) | 6.1±3.9 | 74.5±7.4 | 43.1% |  |
| **ABC-DS,**<br>**total N =**<br>**396** | Controls | 327 (83.4%) | 2.7±0.5 | 45±9.5 | 47.6% | Demographics<br>Diagnosis<br>Physical and<br>neurological<br>examinations<br>Cognitive tests<br>APOE genotype<br>CSF<br>Karyotyping |
|  | MCI | 29 (7.4%) | 2.7±0.5 | 54±5.3 | 43.3% |  |
|  | AD | 40 (10.2%) | 2.9±0.3 | 56±5.4 | 34.5% |  |
| **DIAN, total**<br>**N = 425** | Controls | 304 (71.5%) | 3.1±1.2 | 39.2±10.0 | 57.4% | Demographics<br>Diagnosis<br>Physical and<br>neurological<br>examinations<br>Cognitive tests<br>APOE genotype<br>MRI<br>PET<br>CSF<br>Genetic mutation |
|  | MCI | 45 (10.6%) | 3.2±1.6 | 45.0±9.8 | 71.1% |  |
|  | AD | 76 (17.9%) | 3.5±1.5 | 50.6±7.8 | 46.1% |  |

**<u>TADPOLE</u>**: The Alzheimer’s Disease Prediction Of Longitudinal Evolution (TADPOLE) dataset [18] was drawn from the Alzheimer’s Disease Neuroimaging Initiative (ADNI) [5] and included individuals across the late-onset Alzheimer’s disease spectrum. The dataset comprised N = 1669 subjects, including 502 cognitively normal controls (30.1%), 617 individuals with mild cognitive impairment (37.0%) and 550 with Alzheimer’s disease (33.0%). The data collected included a comprehensive set of biomarkers—neuroimaging (MRI and PET), cerebrospinal fluid (CSF) analytes, cognitive assessments and demographic variables, measured over multiple clinical visits (mean 6.7 visits per participant).

**<u>ABC-DS</u>**: The Alzheimer’s Biomarker Consortium - Down Syndrome study [19] focused on individuals with Down syndrome at high risk for early-onset Alzheimer’s disease due to trisomy 21. The cohort included N = 396 participants with Down syndrome, consisting of N = 327 cognitively normal individuals (83.4%), N = 29 with mild cognitive impairment (7.4%), and N = 40 with Alzheimer’s disease (10.2%). The longitudinal data (mean 2.8 visits per participant) collected included detailed cognitive tests, blood-based biomarkers, and demographic information. We did not have access to neuroimaging measures for this cohort.

**<u>DIAN</u>**: The Dominantly Inherited Alzheimer Network dataset [23] comprised participants carrying autosomal-dominant mutations-in **PSEN1**, **PSEN2**, or **APP**-that lead to familial early-onset AD. This dataset contained N = 425 individuals from families with autosomal dominant Alzheimer’s disease mutations, including N = 304 cognitively normal participants (71.5%), N = 45 with mild cognitive impairment (10.6%) and N = 76 with Alzheimer’s disease (17.9%). This cohort included multimodal biomarkers similar to TADPOLE (neuroimaging, fluid biomarkers, cognitive assessments) but in a genetically defined population with a virtually certain disease trajectory [23] (mean 3.2 visits per participant).

### 1.2 Experimental Setup and Evaluation

We trained and assessed our model on each of the three datasets independently to evaluate performance across diverse Alzheimer’s disease populations. For each dataset, we employed a stratified data splitting strategy to ensure robust and unbiased assessment. We first created a hold-out test set comprising 10% of the subjects from each individual dataset, stratified by age, number of clinical visits, gender, and diagnostic status (CN, MCI and AD), to ensure the test set contained proportionally representative samples compared to the original dataset (see **Supplementary Figures 1, 2 and 3**). The remaining 90% of each dataset was used for model training with 5-fold cross-validation, which helped mitigate potential selection bias and overfitting.

We used all longitudinal data up to each participant’s second-to-last visit to generate month-by-month diagnostic predictions for the subsequent 10-year period. Each participant’s actual diagnosis at their final visit, which occurred at varying points within this window, served as a ground truth to validate the predicted trajectory. This approach evaluated the model’s ability to forecast disease progression across diverse temporal horizons, as subjects’ final visits occurred at different intervals from baseline.

### 1.3 Data Preprocessing and Organization

#### 1.3.1 Missing Data Imputation

A significant challenge in making predictions from longitudinal Alzheimer’s data is the prevalence of missing values, both within individual clinical visits and across the temporal sequence. We employed a K-Nearest Neighbours (KNN) imputation strategy to address this challenge [27]. For baseline clinical visits, we implemented KNN imputation to ensure all participants had complete initial data prior to longitudinal modeling. We identified the K most similar subjects based on available observed biomarkers and utilized their measurements to estimate missing biomarkers for each individual. The similarity between participants was calculated using Euclidean distance in the feature space of available measurements, with K determined through cross-validation on the training data to optimize imputation accuracy while maintaining computational efficiency [27]. For subsequent clinical visits, we applied the same KNN methodology with temporal considerations, where the imputation model incorporated both the cross-sectional information (similarities between participants computed using all available data from baseline through the current timepoint) and longitudinal patterns from the participant’s own measurement history from prior timepoints.

After identifying the K nearest neighbours for a participant with missing values, data imputation was performed using a standard weighted average, where the contribution of each neighbour was inversely proportional to their distance from the target participant:

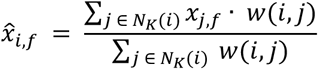

where 𝑥_𝑖,𝑓_ is the imputed value for feature 𝑓 in participant 𝑖, 𝑁_𝐾_(𝑖) represents the set of K nearest neighbours for participant 𝑖 , and 𝑤(𝑖, 𝑗𝑗) is the weight assigned to neighbour 𝑗𝑗 , calculated as 𝑤(𝑖, 𝑗𝑗) = 1/(𝑑(𝑖, 𝑗𝑗) + 𝜀𝜀), with 𝜀𝜀 = 10^−8^ to prevent division by zero when a neighbor has identical feature values to the target participant. This approach ensures that more similar participants contribute more significantly to the imputed values.

#### 1.3.2 Feature Scaling and Normalization

Before model training, all imputed datasets underwent feature scaling and normalization to standardize ranges across heterogeneous biomarkers. Each feature was normalized by subtracting the mean of that feature across subjects and dividing by the standard deviation across subjects calculated from the training data:

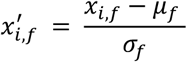

where 𝑥^′^ is the normalized value, and 𝜇_𝑓_ and 𝜎_𝑓_ are the mean and standard deviation of feature 𝑓 across subjects in the training data, respectively.

### 1.4 Time-Aware Feature Attention Mechanism

The model generates individual-specific attention maps revealing biomarker importance across time.

The core innovation of our framework is a multi-dimensional attention mechanism that simultaneously captures both temporal and feature-level importance. This mechanism, illustrated in **Figure 1**, consists of three integrated components that work synergistically to identify the most relevant clinical information across both time and measurement domains.

**Figure 1:**
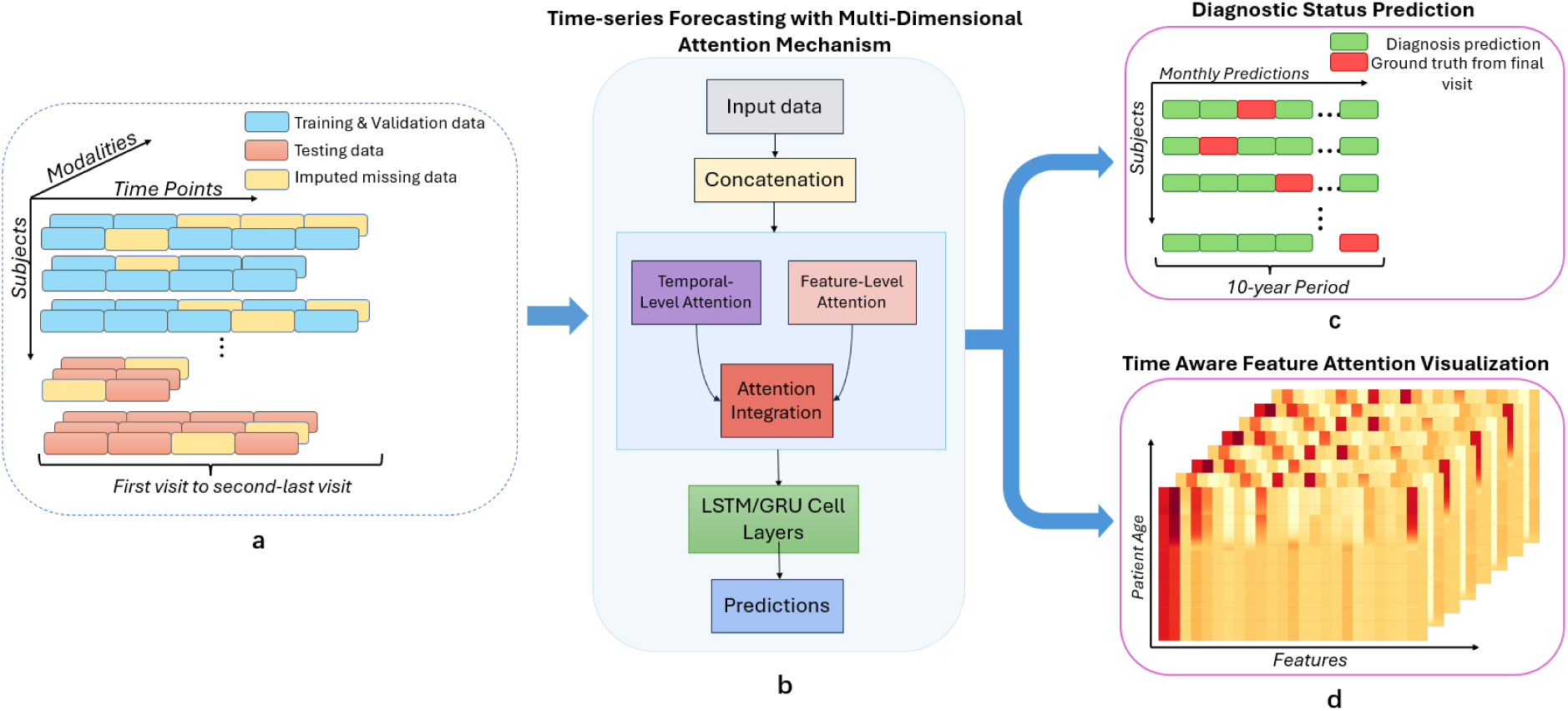
O**v**erview **of the proposed multi-dimensional attention framework for Alzheimer’s disease progression prediction.** (a) Multi-modal longitudinal dataset structure. Each subject has varying numbers of clinical visits over time, with different biomarker modalities collected at each visit. Stratified division into training/validation (blue) and testing (orange) datasets, with missing data imputation (yellow) applied to complete the longitudinal sequences. (b) Multi-dimensional attention framework architecture. The pipeline shows the data flow through temporal attention, feature attention, attention integration and recurrent neural layers (LSTM/GRU) to predict diagnostic status for AD progression. (c) Disease progression forecasting. The model generates month-by-month diagnostic predictions for a 10-year period following each participant’s second-to-last visit. The actual diagnosis at the final visit, occurring at any point within this 10-year window, serves as ground truth for validating the predicted trajectory. (d) Time-aware feature attention visualization.

**Temporal-Level Attention:** This component weighs the importance of different time points in a participant’s clinical history, addressing the fundamental challenge that not all visits carry equal predictive value for disease progression. The temporal attention module incorporates elapsed calendar time between visits as a continuous variable, recognizing that the significance of measurements may decay or evolve non-linearly over time. Mathematically, this temporal weighting is expressed as:

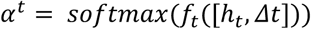

where ℎ_𝑡_ represents the hidden state at time 𝑡, 𝛥𝑡 represents the time elapsed since the previous measurement and 𝑓_𝑡_ is a learned non-linear transformation function implemented as a two-layer neural network with ReLU activation [28]. We employed the SoftMax activation function for temporal attention to create a normalized probability distribution across clinical visits, ensuring the model focused on the most informative visits while maintaining interpretability. This normalized attention distribution enabled the model to identify which clinical visits carried the most prognostic value.

### Feature-Level Attention

This component employed a shared attention mechanism that was learned from all participants during training to identify general patterns of biomarker importance across disease stages. When applied to individual participants, these learned parameters interacted with individual-specific biomarker values to generate personalized attention weights, ensuring that feature importance adapted to each participant’s unique clinical profile while benefiting from population-level insights [29]. The feature attention module employed a learned transformation that mapped input features to attention weights:

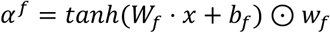

where 𝑊𝑊_𝑓_ and 𝑏_𝑓_ are learnable parameters, 𝑥 is the feature vector, 𝑤_𝑓_ is a feature weight vector, and ⊙ represents element-wise multiplication. We utilized the hyperbolic tangent (𝑡𝑎𝑛ℎ) activation function for feature attention, allowing the model to both emphasize relevant features and suppress misleading ones through its bidirectional output range [30]. Unlike SoftMax, tanh preserves the independent contribution of multiple biomarkers, which is an essential property for Alzheimer’s disease where parallel pathological processes often manifest simultaneously.

**Attention Integration:** The temporal and feature attention mechanisms were integrated through a layer normalization process [31] that ensured stable training dynamics. The integrated attention weights were applied to the input before being processed by the recurrent neural network layers:

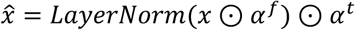

where 𝑥̂ ∈ ℝ^𝑑^ represents the input feature vector at a given time step with 𝑑 dimensions corresponding to different biomarkers, 𝛼^𝑓^ ∈ ℝ^𝑑^ represents the feature attention weights computed through the tanh-based attention mechanism, 𝛼^𝑡^ ∈ ℝ represents the temporal attention weight for the current time step computed through SoftMax normalization, 𝐿𝑎𝑦𝑒𝑟𝑁𝑜𝑟𝑚 denotes the layer normalization operation that standardizes activations across the feature dimension and 𝑥̂ ∈ ℝ^𝑑^ represents the final attention-weighted feature vector. This integration mechanism sequentially applies feature-level attention to identify important biomarkers, normalizes the resulting values to maintain stable gradients, and then applies temporal attention to weight the contribution of different time points in the participant’s disease trajectory [32] [33].

### 1.5 Recurrent Neural Network Architecture

The attention-weighted features from our multi-dimensional attention mechanism were processed through a recurrent neural network that modeled the temporal evolution of biomarkers. We selected recurrent neural networks over alternatives such as transformers primarily due to the variable-length nature of participant data in our datasets. RNNs naturally accommodated this variability without requiring padding or truncation, processing each participant sequence according to its actual length [34].

### 1.6 Implementation Details

Our framework supported multiple RNN variants, including Long Short-Term Memory (LSTM) [13] and Gated Recurrent Unit (GRU) [35]. These gated RNNs selectively remembered long-term dependencies (such as baseline measurements that remain predictive years later) while adaptively incorporating new information when disease state changes occur [14]. The architecture employed dropout regularization applied separately to input and hidden states, multiple stacked RNN layers to capture hierarchical temporal patterns, and residual connections to maintain access to earlier time point information. Our RNN implementation explicitly accounted for irregular intervals between visits by accepting time difference information (𝛥𝑡) as an additional input, allowing the model to adjust its state updates according to the actual elapsed time between measurements.

### 1.7 Transfer Learning and Adaptation

We used transfer learning to leverage the comprehensive biomarker patterns learned from the TADPOLE dataset when adapting to more specialized and data-limited populations represented by ABC-DS and DIAN [36]. Transfer learning selectively preserved knowledge in specific components of our neural architecture [37]. We froze the feature attention mechanism and first recurrent layer, as these components captured fundamental disease progression patterns and biomarker relationships. Meanwhile, subsequent recurrent layers and output projection layers remained trainable, allowing adaptation to population-specific characteristics.

### 1.8 Training Process and Regularization

The model was trained using an early stopping strategy [38] [39] with patience and minimum improvement thresholds to prevent overfitting while ensuring optimal performance. Specifically, we monitored the cross-entropy loss on validation data during training and halted the process when no improvement exceeding a predefined threshold (0.001) was observed for 10 consecutive epochs [38]. This approach automatically determined the optimal training duration for each dataset. After validation performance plateaued, we saved the model weights that achieved the best validation performance. The training process employed cross-entropy loss for diagnostic classification. We implemented learning rate scheduling with step decay to improve convergence, gradually reducing the learning rate when performance plateaued. The learning rate was initialized at 0.001 and reduced by a factor of 0.3 every 5 epochs when validation performance showed no improvement [40]. We also applied an L2 regularization term with a coefficient of 10^−5^ to all model parameters. This regularization approach prevented individual weights from becoming excessively large, effectively constraining the model’s complexity and reducing the risk of overfitting to training data peculiarities that might not generalize to new populations [41] [42].

### 1.9 Evaluation Metrics

The primary evaluation metric we employed was the multi-class Area Under the ROC Curve (mAUC), to assess how accurately our model predicted a participant’s future diagnostic state (CN, MCI, or AD) based on their longitudinal biomarker history, in a way that was robust to class imbalance across the three diagnostic categories.

The mAUC was calculated by averaging the AUC values for all possible pairwise class comparisons:

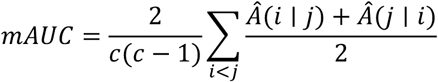

Where 𝐴^(𝑖 ∣ 𝑗𝑗) represents the probability that a randomly drawn member of class 𝑗𝑗 will have a lower estimated probability of belonging to class i than a randomly drawn member of class 𝑖, and c is the total number of classes. This formulation ensures that the metric performed consistently regardless of class imbalance in the dataset and did not require specification of classification thresholds or misclassification costs.

## Results

### 2.1 Performance Comparison

**Table 2** presents the models’ performance across the TADPOLE, ABC-DS and DIAN datasets.

**Table 2.** Performance (mAUC values) of Alzheimer’s disease progression prediction models across TADPOLE, ABC-DS, and DIAN datasets. Models include Support Vector Machine (SVM), Gated Recurrent Unit (GRU), Long Short-Term Memory (LSTM), Multi-Dimensional Attention GRU (our proposed model), and TADPOLE-Pretrained Multi-Dimensional Attention GRU (transfer learning approach). Higher mAUC values (closer to 1) indicate better performance, with 0.5 representing chance level. Bold values indicate the best performance for each dataset.

| *Model* | *TADPOLE* |  | *ABC-DS* |  | *DIAN* |  |
| --- | --- | --- | --- | --- | --- | --- |
|  | Validation | Test | Validation | Test | Validation | Test |
| *SVM* | 0.682 | 0.665 | 0.583 | 0.613 | 0.704 | 0.722 |
| *GRU* | 0.771 | 0.677 | 0.629 | 0.637 | 0.783 | 0.767 |
| *LSTM* | 0.764 | 0.685 | 0.618 | 0.651 | 0.779 | 0.767 |
| *Multi-Dimensional Attention GRU* | **0.856** | **0.793** | 0.642 | 0.680 | **0.814** | **0.902** |
| *TADPOLE Pretrained Multi-Dimensional Attention GRU* | N/A | N/A | **0.779** | **0.771** | 0.810 | **0.902** |

For the TADPOLE dataset, the multi-dimensional attention GRU achieved mAUC values of 0.856 (validation) and 0.793 (test), outperforming the standard GRU (0.771/0.677), LSTM (0.764/0.685), and baseline SVM (0.682/0.665). The validation-test gap for the attention model was 0.063, compared to 0.094 for GRU and 0.079 for LSTM, suggesting better generalization to unseen participants.

The same performance pattern was observed in the ABC-DS and DIAN cohorts. In ABC-DS (without transfer learning), the multi-dimensional attention GRU achieved 0.642/0.680 compared to the standard GRU’s performance of 0.629/0.637. For DIAN, the attention model reached 0.814/0.902 versus 0.783/0.767 for the standard GRU. Across all three datasets, the performance ranking remained consistent: multi-dimensional attention GRU achieved the highest mAUC values, followed by the LSTM and standard GRU, with the baseline SVM showing the lowest performance.

### 2.2 Transfer learning from TADPOLE dataset

Transfer learning from TADPOLE improved the ABC-DS validation performance by 21.2% (from 0.642 to 0.779) and test performance by 13.4% (from 0.680 to 0.771). We note that the ABC-DS dataset did not include the neuroimaging features present in TADPOLE, suggesting that pre-training on neuroimaging data can improve prediction accuracy on data where no neuroimaging is available, likely due to disease mechanisms that transcend etiological boundaries and data modalities.

The prediction metrics in the DIAN dataset showed minimal change with transfer learning, with validation performance decreasing by 0.004 and test performance remaining unchanged at 0.902. This suggests that populations with deterministic mutations and relatively comprehensive multimodal data may not benefit from knowledge transfer from sporadic AD cohorts.

### 2.3 Stratified Performance Analysis

Stratified analysis of our multi-dimensional attention GRU model on the TADPOLE cohort revealed how participant characteristics influenced prediction accuracy. The strongest predictor was the number of longitudinal assessment points: participants with six or more visits achieved an average mAUC of 0.917, compared to 0.826 for 4-5 visits and 0.808 for 2-3 visits, confirming-as expected-that temporal attention mechanisms benefit from richer longitudinal information. Age showed an inverse relationship with mean accuracy, declining from 0.876 in participants under 65 to 0.826 (ages 65-75) and 0.813 (over 75), likely reflecting the challenge of distinguishing pathological decline from normal aging-related changes in older participants.

The interval between the penultimate and final assessments had limited impact on prediction accuracy (0-6 months: 0.803; 6-18 months: 0.821; >18 months: 0.786), suggesting the model maintains robustness across different prediction horizons. Similarly, demographic factors showed negligible effects: the model achieved nearly identical performance on female and male participants (0.820 vs 0.819). While the TADPOLE cohort lacks significant racial diversity (predominantly White at 83.8%), analysis of the available data showed limited performance variation across groups: Asian (mAUC = 0.831, N = 78), More than one race (mAUC = 0.824, N = 71), White (mAUC = 0.820, N = 1394), apart from somewhat reduced accuracy for Black participants (mAUC = 0.787, N = 118). These findings suggest the model mostly captures disease patterns rather than demographic confounds, although further work in more diverse samples including greater numbers of Black participants will be important.

### 2.4 Feature Attention Analysis – TADPOLE

Figure 2a illustrates a personalized attention heatmap for a representative subject from the TADPOLE dataset, demonstrating how feature importance evolves uniquely across their individual disease trajectory from age 74 to 83. We computed the distribution of personalized feature attention weights for the top 10 features plus APOE4 across all TADPOLE participants, see Figure 2b. We observed substantial heterogeneity in attention patterns, with hippocampal volume and cognitive measures showing wide variances, suggesting diverse progression phenotypes even within sporadic Alzheimer’s disease. Diagnosis and age demonstrated the highest median attention values (0.086 and 0.077 respectively) with relatively tight distributions (coefficient of variation < 0.20), confirming their role as anchoring features across all participants.

**Figure 2:**
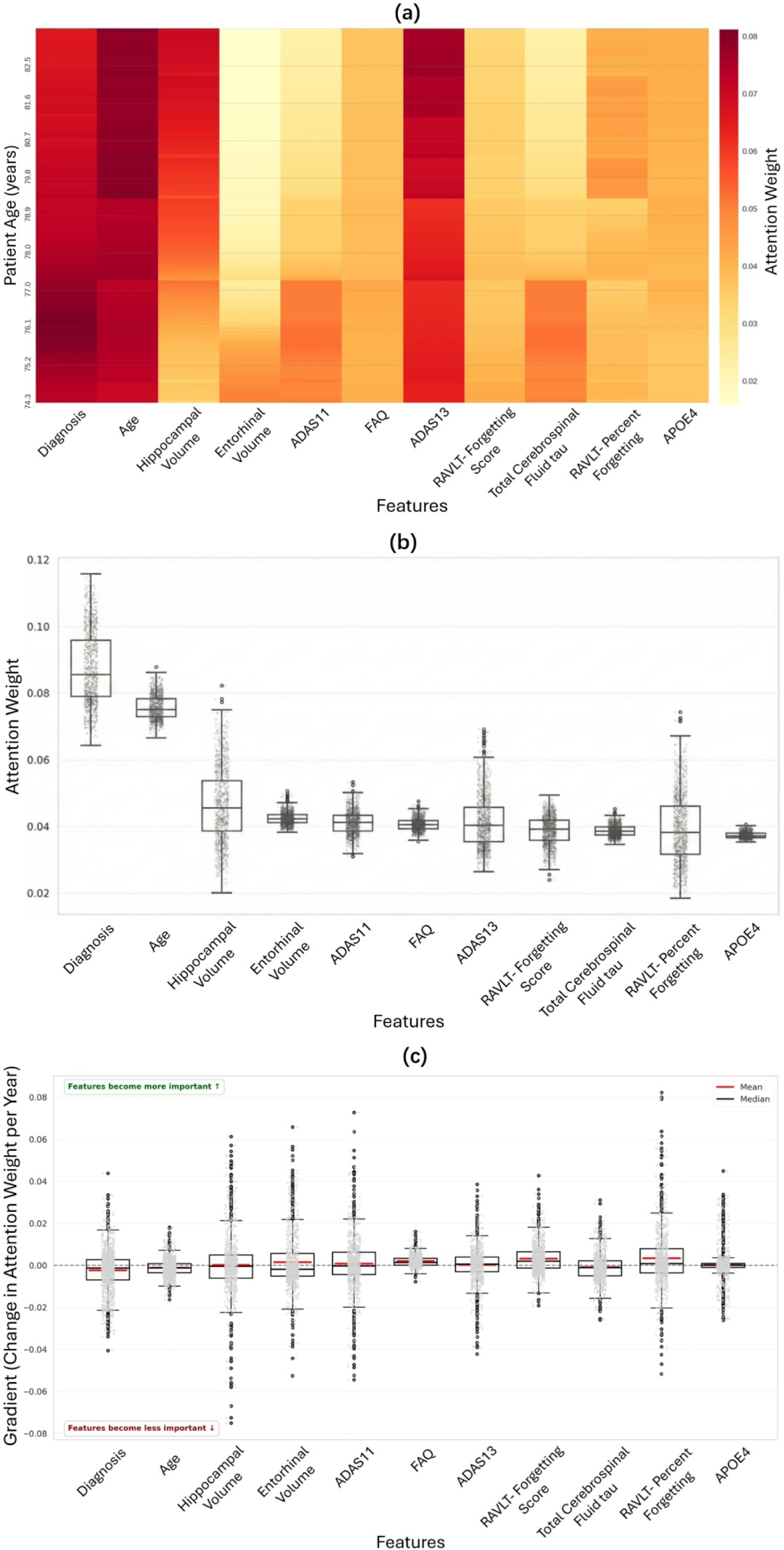
F**e**ature **attention analysis for the TADPOLE cohort. (a) Individual-Specific Attention Patterns of a TADPOLE Participant:** This example individual participant’s attention map showed high personalized weights for diagnosis and age throughout their specific disease trajectory from age 74-83. Cognitive measures (Alzheimer’s Disease Assessment Scale-Cognitive Subscale 13 items (ADAS13)) and structural markers (Hippocampal Volume) showed increasing importance with age. The participant was diagnosed as MCI at age 81. **(b) Distribution of Personalized Feature Attention Weights Across TADPOLE Cohort:** Diagnosis and age dominated with highest attention weights, while cognitive (Alzheimer’s Disease Assessment Scale-Cognitive Subscale 11 items (ADAS11), ADAS13, Functional Activities Questionnaire (FAQ), Rey Auditory Verbal Learning Test (RAVLT)) and structural (Hippocampal and Entorhinal Volume) markers showed moderate importance with greater inter-participant variability. APOE4 demonstrated lowest but consistent attention across participants, aligning with its role in disease initiation rather than progression monitoring. **(c) Temporal Gradients of Feature Importance Across Disease Progression of TADPOLE Cohort:** Feature importance shifted from molecular markers (Total Cerebrospinal Fluid tau) to cognitive-functional measures (RAVLT, FAQ) and structural markers (Entorhinal Volume) as Alzheimer’s disease progressed, capturing the evolution from biochemical changes to clinical symptoms.

Finally, Figure 2c presents the temporal gradients of feature importance, revealing a fundamental shift in biomarker relevance during AD progression. Gradients were calculated as the change in attention weight per year of follow-up, capturing whether a feature’s importance increased or decreased over time. The gradient analysis uncovered a critical transition from molecular to clinical markers as the disease advances. Notably, the ADNI total tau biomarker exhibited a negative gradient (reduced feature importance over time), potentially in-line with recent evidence that CSF total tau biomarkers can plateau in advanced disease stages due to extensive neuronal loss reducing the available substrate for tau release [44] [45] [43], and longitudinal studies showing tau accumulation rates diminish as available binding sites become saturated [47] [48]. Conversely, cognitive and functional measures demonstrated positive gradients, indicating their importance increased over time. The Rey Auditory Verbal Learning Test score (a measure of memory consolidation failure), the Alzheimer’s Disease Assessment Scale-Cognitive Subscale (a measure of cognitive functional decline), and the Functional Activities Questionnaire (a measure of instrumental activities) showed stage-dependent predictive importance. Memory measures peaked in early stages, while functional measures became critical in later progression. This pattern captured the biological reality that as molecular pathology plateaus, clinical manifestations become the primary drivers of diagnostic transitions. The model successfully identified this shift from upstream molecular processes to downstream clinical consequences.

### 2.5 Feature Attention Analysis - ABC-DS and DIAN

Figures 3 **and 4** present the distributions of feature importance and feature importance gradients for the ABC-DS and DIAN cohorts.

**Figure 3:**
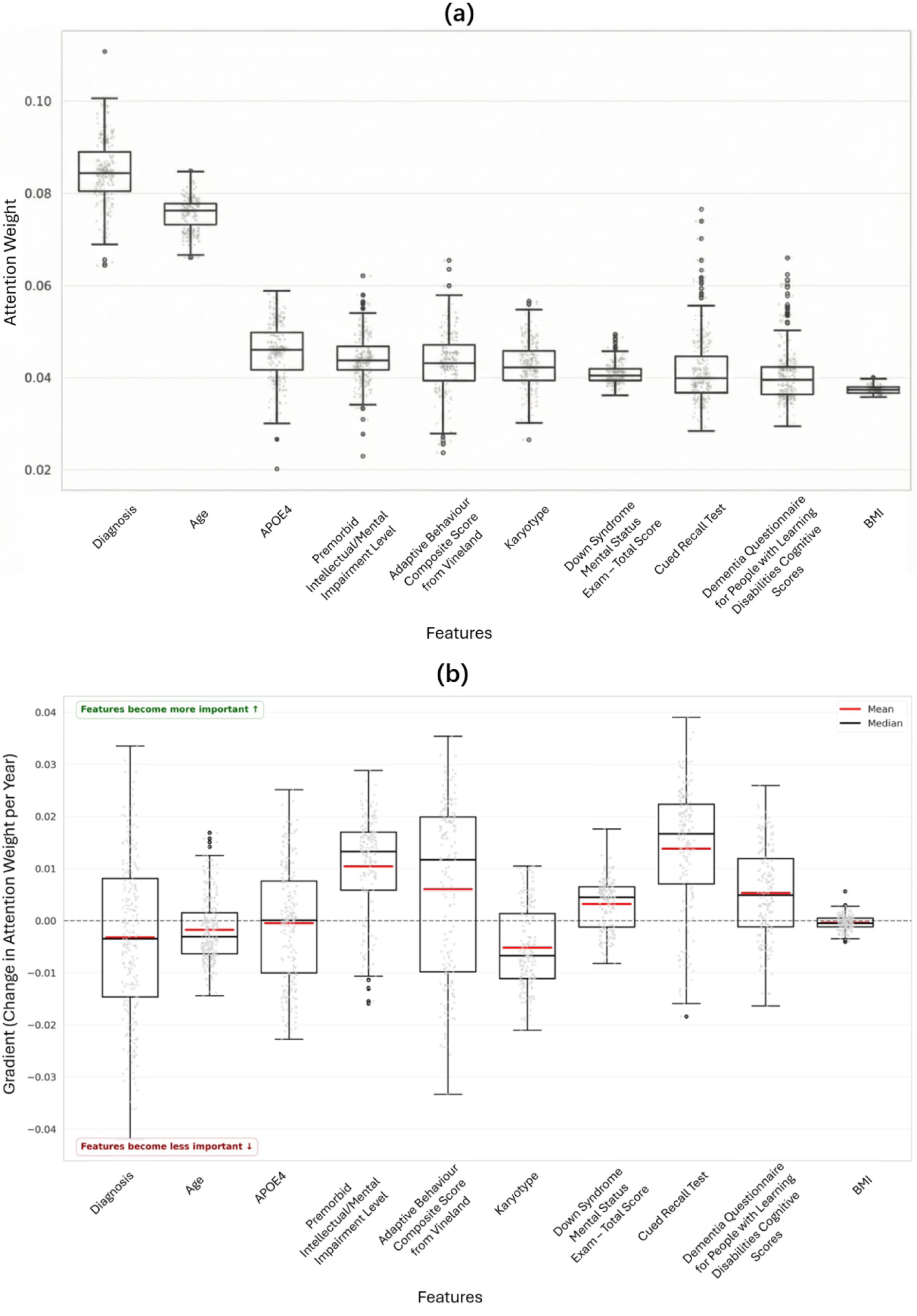
F**e**ature **attention analysis for the ABC-DS cohort. (a) Distribution of Personalized Feature Attention Weights Across ABC-DS Cohort:** APOE4 showed high median attention across participants. Memory scores, functional measures and Karyotype received elevated attention weights. **(b) Temporal Gradients of Feature Importance Across Disease Progression of ABC-DS Cohort:** Memory and functional measures (Cued Recall Test, Premorbid Intellectual/Mental Impairment Level, and Adaptive Behaviour Composite Score from Vineland) displayed the strongest positive gradients, while Karyotype exhibited the most negative gradient, followed by Age and Diagnosis.

**Figure 4:**
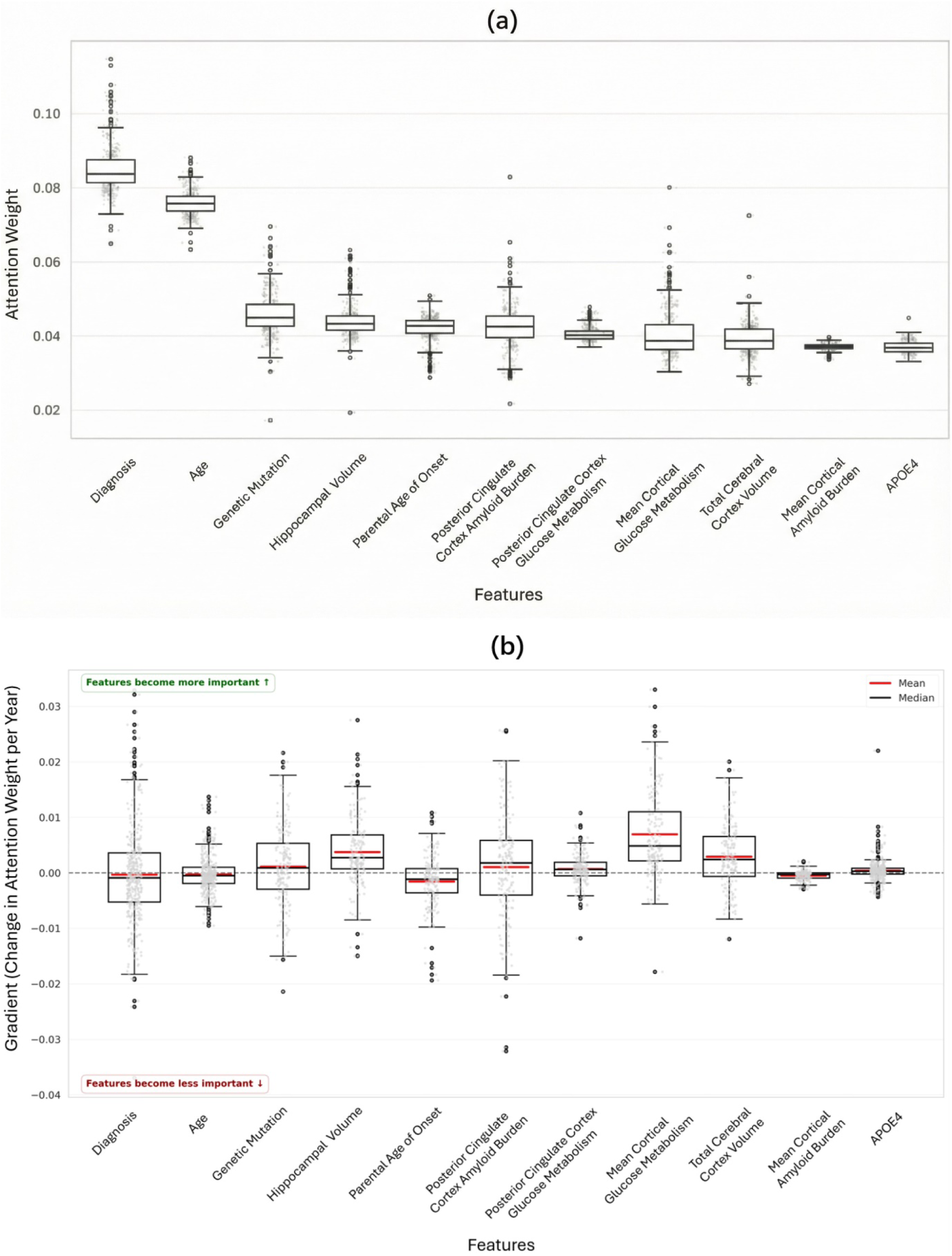
F**e**ature **attention analysis for the DIAN cohort. (a) Distribution of Personalized Feature Attention Weights Across DIAN Cohort:** Genetic markers (Genetic Mutation, Parental Age of Onset) showed high attention weights. Neuroimaging measures (Hippocampal Volume, Posterior Cingulate Cortex Amyloid Burden, Posterior Cingulate Cortex Glucose Metabolism, Mean Cortical Glucose Metabolism, Total Cerebral Cortex Volume, and Mean Cortical Amyloid Burden) demonstrated elevated attention compared to cognitive measures in TADPOLE and ABC-DS cohorts. **(b) Temporal Gradients of Feature Importance Across DIAN Disease Progression:** Parental Age of Onset exhibited strongest negative gradient. Neuroimaging measures showed positive gradient or near-zero patterns.

#### ABC-DS Adaptations

In the absence of neuroimaging data, the model adapted by elevating attention to available clinical markers, see Figure 3a and b. APOE4 showed a high median attention weight (0.045) with a wide variance, indicating heterogeneous genetic influence. Clinical measures including blood pressure and daily function scores received elevated attention weights. The temporal gradient analysis showed memory and functional measures (Cued Recall Test, Premorbid Intellectual/Mental Impairment Level, and Adaptive Behaviour Composite Score from Vineland) displaying the strongest positive gradients (median gradient >0.01). Age demonstrated a stronger negative gradient compared to TADPOLE patterns. This reflects the deterministic nature of Alzheimer’s pathology in Down syndrome, where virtually all individuals develop amyloid pathology by age 40 due to APP gene triplication, making chronological age less predictive once individuals enter the typical onset window.

Diagnosis exhibited a negative median gradient with high variance across participants. Standard diagnostic categories provide inconsistent predictive value in Down syndrome due to pre-existing intellectual disabilities of varying severity, which obscure clear diagnostic transitions that would typically indicate progression. The high variance reflects substantial individual differences, with some participants showing clear diagnostic progression while others are already impaired at baseline, making diagnosis unreliable as a universal predictor in this population. Karyotype showed the strongest negative gradient, reflecting its diminishing utility as a predictor once individuals enter the deterministic disease window.

#### DIAN Characteristics

Genetic markers (Mutation) demonstrated the highest attention weights, with mutation status and parental age of onset showing median attention >0.04, see Figure 4a and b. PIB PET measures (Posterior Cingulate Cortex Amyloid Burden and Mean Cortical Amyloid Burden) displayed elevated importance (median: 0.043 and 0.038 respectively), reflecting strong amyloid pathology signatures. Several neuroimaging measures (Hippocampal Volume, Mean Cortical Glucose Metabolism and Total Cerebral Cortex Volume) showed strong positive gradients (median gradient >0.025), reflecting the importance of neuroimaging data in predicting and modeling the progression of AD.

## Discussion

### 3.1 Model Performance Across Populations

Our multi-dimensional attention framework achieved substantial performance improvements over baseline models. This consistent enhancement demonstrated that attention mechanisms captured clinically meaningful patterns beyond conventional recurrent architectures, aligning with recent findings in medical time series analysis [28]. The largest improvement in performance occurred in the DIAN cohort, where the independently trained multi-dimensional attention model achieved a 0.135 mAUC increase over the standard GRU model (0.902 vs 0.767). This improvement highlights how the attention mechanism effectively captured the predictable progression patterns created by deterministic mutations [23]. Conversely, the model showed the smallest gain over the standard GRU in the ABC-DS dataset without transfer learning (0.680 vs 0.637). This more modest increase reflects the challenges posed by limited biomarker availability and fewer clinical visits per participant in this dataset [19] [2].

### 3.2 Transfer Learning

#### 3.2.1 Knowledge Transfer Across Population Barriers

We observed a significant improvement in model performance in the ABC-DS cohort when the model was first trained on the TADPOLE cohort: validation mAUC improved from 0.642 to 0.779 (21.2%) and test mAUC from 0.680 to 0.771 (13.4%). This suggested our model captured fundamental disease mechanisms that are shared across distinct AD etiologies. The efficacy of transfer learning for the ABC-DS cohort, which lacks the neuroimaging data of the TADPOLE training set, illustrated that the learned progression patterns are robust to variations in both disease etiology and data modality.

Conversely, in the DIAN cohort pre-training on the TADPOLE dataset had very little impact on the model’s already high predictive performance, with test mAUC remaining unchanged at 0.902 and validation mAUC showing only a marginal decrease from 0.814 to 0.810. The absence of detrimental interference here suggests that the disease progression patterns learned from the sporadic cohort are fundamentally compatible with the pathway observed in the autosomal dominant cohort. This compatibility reinforces the concept of a shared underlying neurodegenerative process, while also demonstrating that our framework can achieve high accuracy independently when comprehensive multimodal data is available.

#### 3.2.2 Biological Significance

These results align with the biological understanding that while the initial causes of Alzheimer’s disease may differ between populations (sporadic, chromosomal, or autosomal dominant), certain downstream pathological processes share common pathways [47] [48]. Our model’s ability to transfer knowledge between three distinct populations provided computational evidence supporting the concept that while AD initiation mechanisms differ, the neurodegenerative cascade followed conserved pathways involving tau propagation, synaptic dysfunction, and neuronal loss.

#### 3.2.3 Clinical Utility of Transfer Learning

The performance enhancements achieved through transfer learning also have significant clinical implications. For populations with Down syndrome, where comprehensive biomarker collection presents logistical and practical challenges, the proposed methodology facilitated more accurate disease progression prediction and staging despite not having access to neuroimaging data from ABC-DS. This advancement has direct relevance for clinical trial design, including better participant stratification without neuroimaging, more accurate sample size estimations, and improved identification of rapid progressors for enrichment strategies. These capabilities might also enhance longitudinal patient monitoring in specialized populations where imaging is impractical, economically expensive or burdensome for patients. Furthermore, the successful transfer learning from TADPOLE to ABC-DS demonstrates that the model can learn generalizable disease principles while retaining population-specific flexibility [36], suggesting potential for a universal Alzheimer’s progression model that adapts to available data rather than requiring full retraining for each cohort [49].

### 3.3 Feature Importance

#### 3.3.1 Biological Significance

The model identified three distinct biomarker tiers that emerged without a predefined hierarchy: anchoring features (diagnosis, age) with consistently high attention throughout disease progression; progression markers (cognitive/structural measures) with moderate attention and high variability reflecting individual differences; and risk factors (APOE4) with high baseline importance. This stratification aligned with established biology where current state indicators remained consistently important, progression markers tracked ongoing neurodegeneration [26], and genetic factors were crucial for determining initial disease risk but less informative for monitoring ongoing disease progression because they are static and do not change over time [50]. The population-specific gradient signatures further validated that our model captured genuine biological heterogeneity, with each cohort displaying distinct temporal dynamics that aligned with their known pathophysiological characteristics rather than representing computational artifacts or overfitting patterns.

Population-specific patterns validated known pathophysiological differences. In ABC-DS, the prominence of APOE4 as the highly weighted feature with substantial inter-participant variance reflected the unique interaction between APOE genotype and trisomy 21, where the triplicated APP gene on chromosome 21 may modulate APOE’s effects differently compared to sporadic Alzheimer’s disease. DIAN’s genetic marker dominance captured the 72% variance in symptom onset explained by parental age of onset, with negative gradients reflecting diminishing importance once phenotype emerges [20].

Our temporal gradient analysis revealed that biological factor importance is fundamentally stage dependent. The tau biomarkers showed decreasing predictive value in late disease due to plateau effects, while cognitive measures demonstrated increasing importance as disease advanced. This stage-dependency has critical implications for clinical monitoring strategies.

#### 3.3.2 Clinical Implications of Personalized Attention Patterns

The individual-specific attention mechanisms proposed here could enable precision monitoring strategies tailored to individual disease trajectories. The temporal gradient analysis suggested that participants showing increasing importance for certain biomarkers over time may benefit from more frequent assessments of those markers, while stable or decreasing gradients could indicate opportunities to reduce monitoring burden. This data-driven approach might optimize both clinical resources and patient burden, consistent with precision medicine frameworks [20] [1].

The negative tau gradient coupled with positive cognitive measure gradients suggested that early-stage monitoring might best prioritize CSF biomarkers [51], while late-stage monitoring might better emphasize cognitive assessments [52]. This stage-dependent strategy reflects the plateau phenomenon observed in CSF total tau during advanced disease stages [46], where molecular biomarkers reach ceiling effects while cognitive measures continue tracking disease progression, potentially optimizing both resource allocation and diagnostic accuracy throughout the disease continuum. Potential population-specific strategies also emerged from the attention patterns. TADPOLE participants might be best suited to balanced multimodal monitoring [18], ABC-DS participants showed high attention to APOE4 status and functional measures [19], and DIAN participants needed early genetic characterization transitioning to intensive multimodal monitoring near expected onset age [23]. Nonetheless, we note that for all datasets and most features there was high heterogeneity in feature importance between participants, potentially suggesting the importance of tailoring these strategies to specific individuals.

### 3.4 Limitations

While our multi-dimensional attention framework demonstrates strong performance across diverse AD populations, several limitations warrant consideration:

**Population Representativeness**: The three cohorts, while diverse in etiology, do not represent the full spectrum of AD. TADPOLE participants are predominantly well-educated volunteers, ABC-DS represents a specific genetic condition, and DIAN includes only rare mutation carriers. Generalization to community-based populations requires further validation. Furthermore, while the model showed similar performance across most demographic groups, reduced accuracy was observed for Black participants (mAUC = 0.787 compared to 0.820 for White participants). This finding requires cautious interpretation given the limited sample size (N = 118) and warrants further investigation in more diverse cohorts.

**Attention Interpretability**: While our attention mechanism provides unique individual-specific insights into biomarker importance, it shares the interpretability challenges common to deep learning models [53]. In particular, the attention mechanism cannot disambiguate between correlated features. When biomarkers are highly correlated, the model may assign high attention to one while ignoring others that are equally biologically important. For example, if hippocampal volume and entorhinal cortex thickness are correlated, the model might attend only to hippocampal volume even though both reflect the same underlying neurodegenerative process. Therefore, low attention does not imply biological irrelevance, as the feature’s information may be captured through its correlated partners. High attention to a biomarker also does not specify how clinicians should act on this information. For instance, high attention to hippocampal volume tells us this feature is predictive for a given individual, but does not indicate whether the participant’s values are clinically concerning or what actions clinicians should take in response. Future work should focus on developing decision support frameworks that translate attention patterns into actionable clinical recommendations through collaboration with clinical experts.

**Gradient Stability**: The high variance in temporal gradients, particularly in ABC-DS and DIAN, suggests that longer follow-up periods may be needed to establish stable importance trajectories for some biomarkers.

### 3.5 Conclusions

Multi-dimensional attention mechanisms successfully modelled Alzheimer’s disease progression across diverse populations with higher accuracy than conventional machine learning approaches. Importantly, pre-training on a general late-onset population sample substantially improved prediction accuracy in a more data-limited, early-onset genetic cohort, suggesting shared underlying disease mechanisms and demonstrating that knowledge from well-characterized cohorts can enhance prediction in understudied populations, including populations where neuroimaging data may not be available. Our framework also enables the generation of personalized attention patterns, revealing both universal disease principles and population-specific adaptations that reflect distinct genetic architectures. Ultimately, these results position our approach not as a black-box predictor, but as a hypothesis-generating tool that complements rather than replaces clinical expertise in precision medicine applications.

## Data Availability

The TADPOLE dataset is available through the Alzheimer's Disease Neuroimaging Initiative (ADNI) at adni.loni.usc.edu. The ABC-DS dataset is available through the National Institute on Aging at https://www.nia.nih.gov/research/abc-ds. The DIAN dataset is available through the Dominantly Inherited Alzheimer Network at https://dian.wustl.edu/. Access to these datasets requires application approval due to participant privacy protections.

https://adni.loni.usc.edu/

https://www.nia.nih.gov/research/abc-ds

https://dian.wustl.edu/

## Acknowledgements

Data used in preparation of this article was obtained from the Alzheimer’s Disease Neuroimaging Initiative (ADNI) database, the Alzheimer’s Biomarker Consortium–Down Syndrome (ABC-DS) database, and the Dominantly Inherited Alzheimer Network (DIAN) database. As such, the investigators within ADNI, ABC-DS and DIAN contributed to the design and implementation of ADNI, ABC-DS and DIAN and/or provided data but did not participate in analysis or writing of this report. A complete listing of ADNI investigators can be found at: http://adni.loni.usc.edu/wp-content/uploads/how_to_apply/ADNI_Acknowledgement_List.pdf

A complete list of ABC-DS investigators can be found at: https://www.nia.nih.gov/research/abc-ds#study-sites-and-investigators

A complete list of DIAN investigators can be found at: https://dian.wustl.edu/for-investigators/diantu-investigator-resources/dian-tu-study-team/

## Author Contributions

Z.S. built the machine learning models and performed all data analyses. S.E.M. and S.Z. supervised the work. All authors provided substantial contributions to the discussion of content, wrote the article and reviewed/edited the manuscript before submission.

## Competing Interests

The authors declare no competing interests.

## Notes

### Competing Interest Statement

The authors have declared no competing interest.

### Funding Statement

No specific funding was received for this work.

### Author Declarations

This study used de-identified data from three existing research consortia. The Alzheimer's Disease Neuroimaging Initiative (ADNI) data were accessed under ADNI's data use agreement, with original ethics approval from institutional review boards at each ADNI site. The Alzheimer's Biomarker Consortium - Down Syndrome (ABC-DS) data were accessed under ABC-DS's data use agreement, with original ethics approval from institutional review boards at participating ABC-DS sites. The Dominantly Inherited Alzheimer Network (DIAN) data were accessed under DIAN's data use agreement at Washington University in St. Louis, with original ethics approval from institutional review boards at participating DIAN sites. The current study involved secondary analysis of existing de-identified data.

