## Supplementary Information for "Multi-dimensional attention framework for personalized Alzheimer’s disease progression prediction across sporadic and genetic risk cohorts"

### Supplementary Methods

(a)

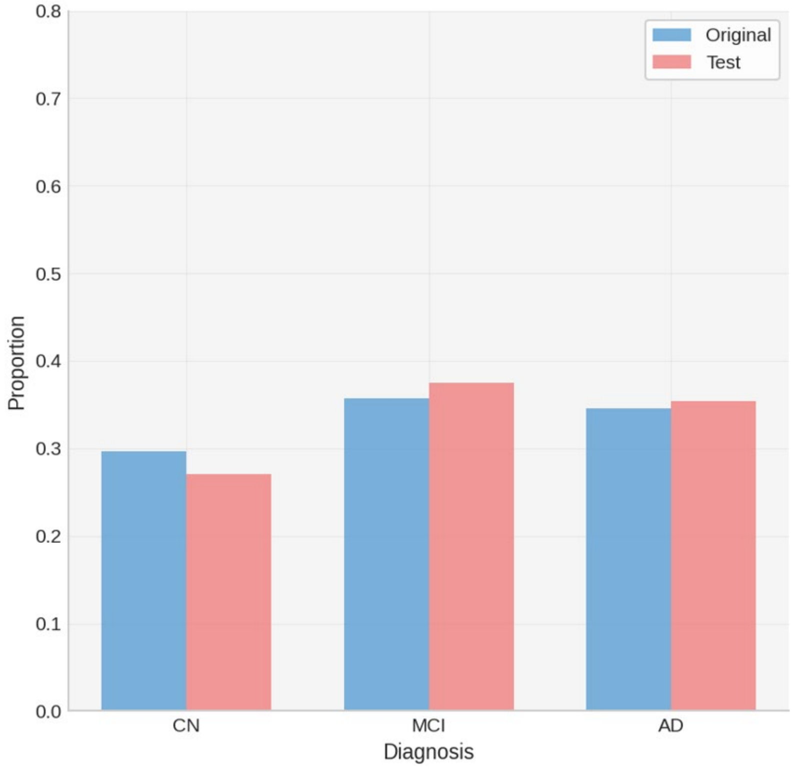


(b)

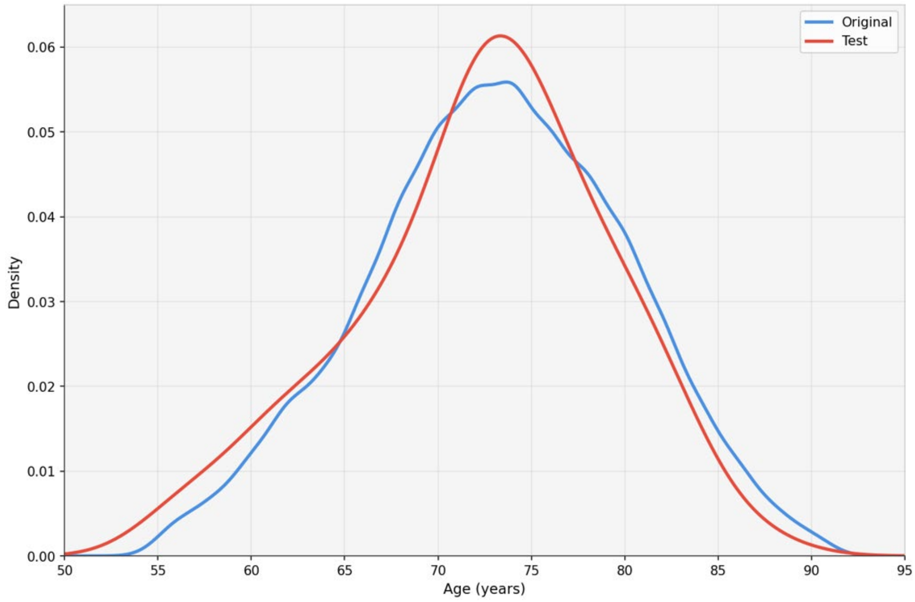

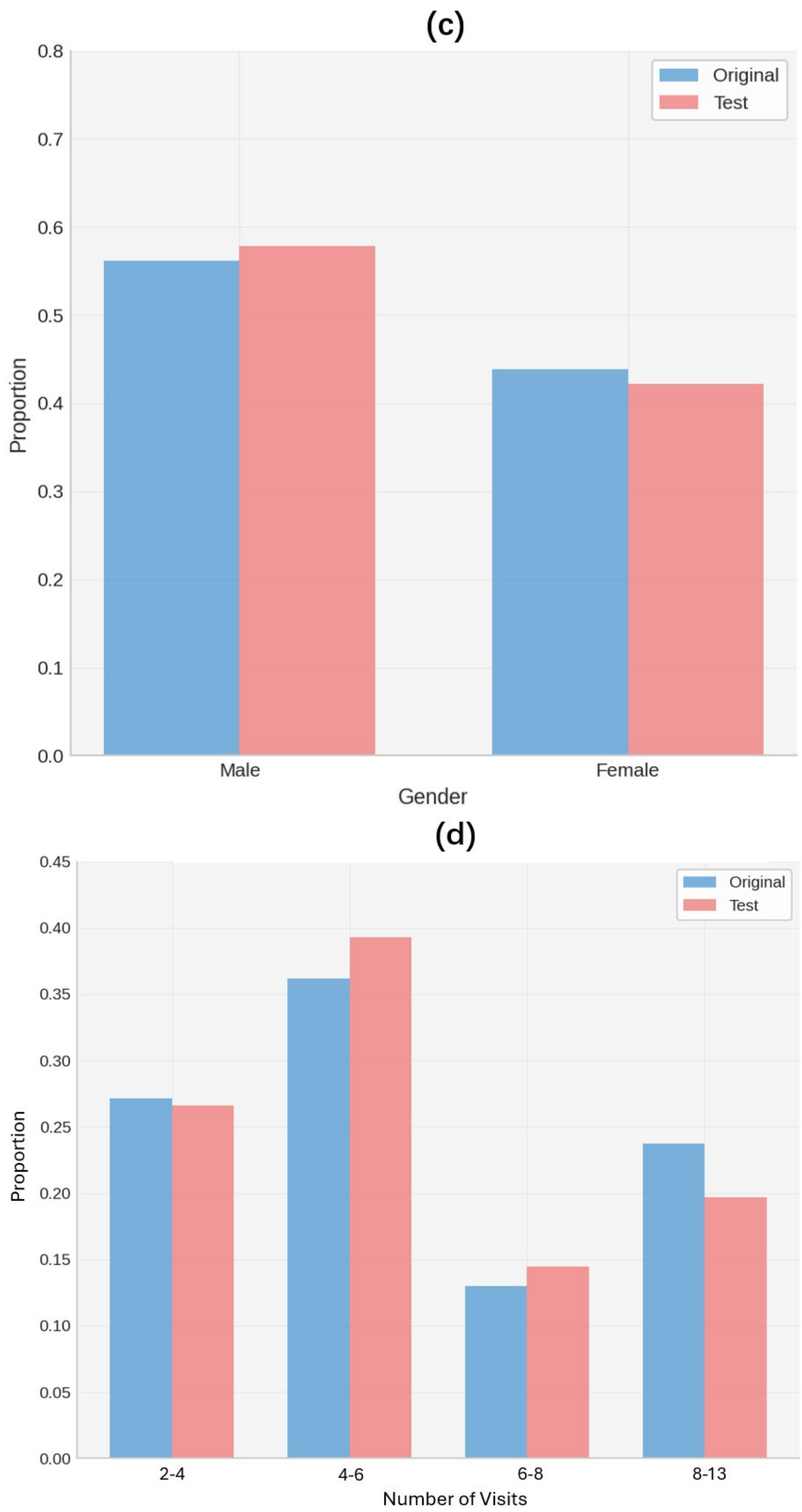


Supplementary Figure 1. Stratified data splitting strategy for TADPOLE dataset. (a) Diagnosis distribution in original and test sets. (b) Age distribution in original and test sets. (c) Gender distribution in original and test sets. (d) Number of clinical visits distribution in original and test sets. The distribution plots showed that the stratified data splitting strategy considered multiple factors to retain the almost same characteristics between the original dataset and the hold-out testing set, ensuring the proposed framework was evaluated fairly.

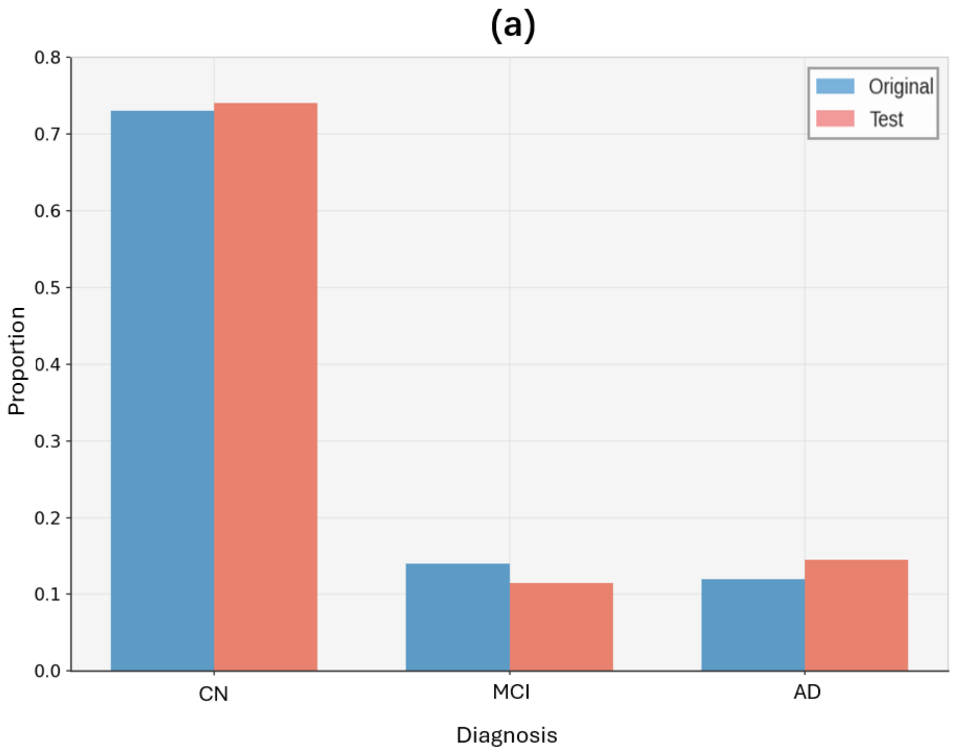

(a)
Original
Test
0.8
0.7
0.6
0.5
0.4
0.3
0.2
0.1
0.0
Proportion
CN
MCI
AD
Diagnosis

(b)

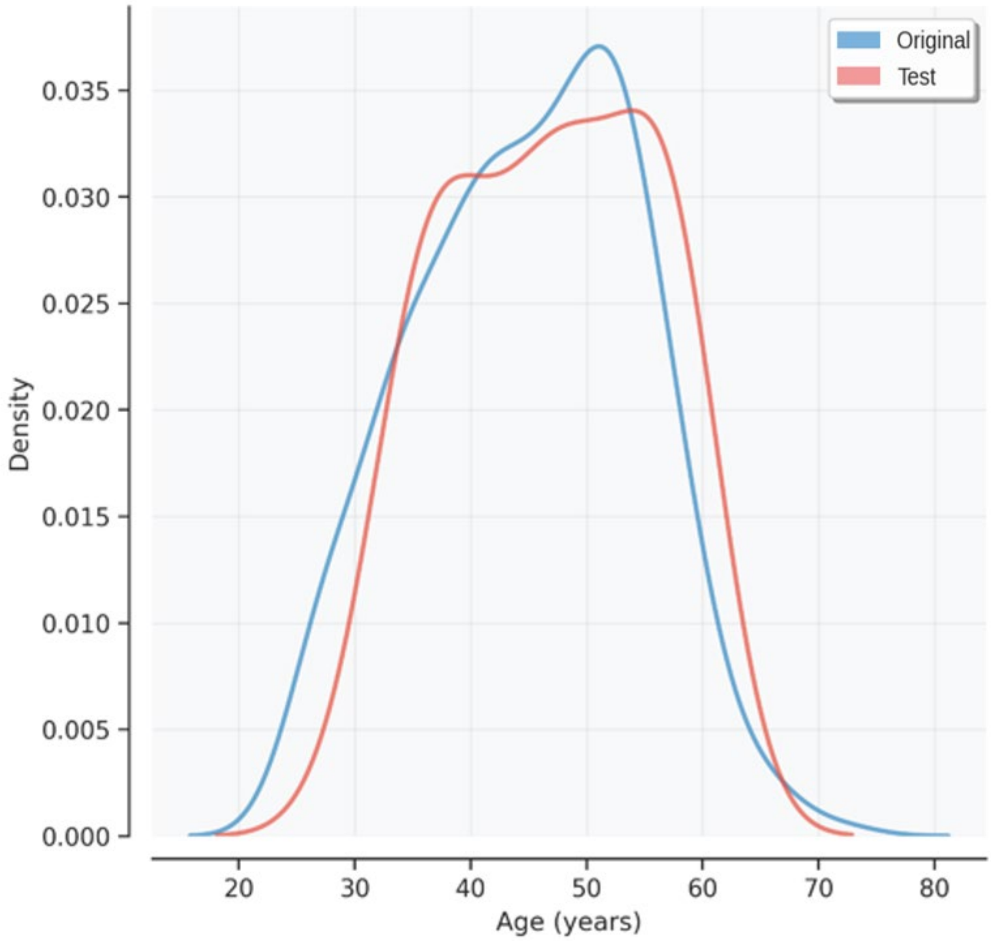
Original
Test
Density
0.035
0.030
0.025
0.020
0.015
0.010
0.005
0.000
20
30
40
50
60
70
80
Age (years)

(c)

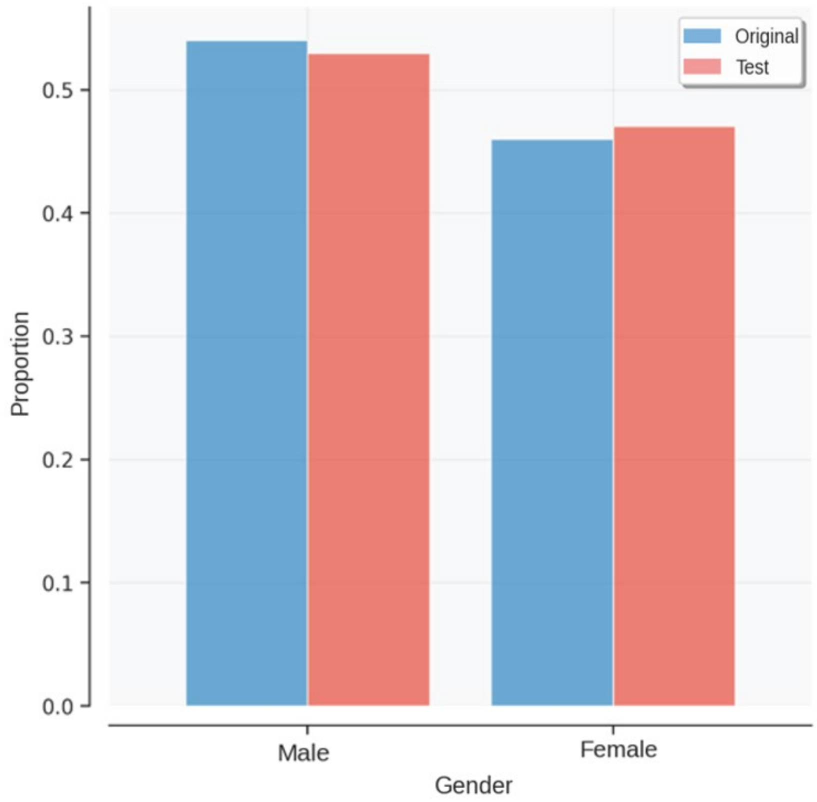


(d)

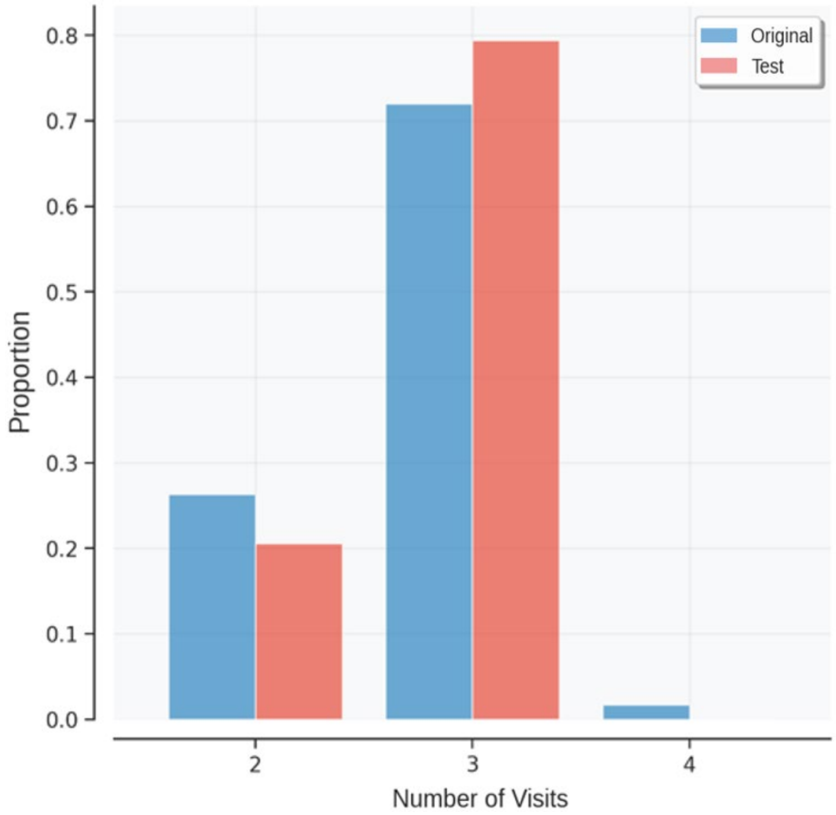


Supplementary Figure 2. Stratified data splitting strategy for ABC-DS dataset. (a) Diagnosis distribution in original and test sets. (b) Age distribution in original and test sets. (c) Gender distribution in original and test sets. (d) Number of clinical visits distribution in original and test sets.

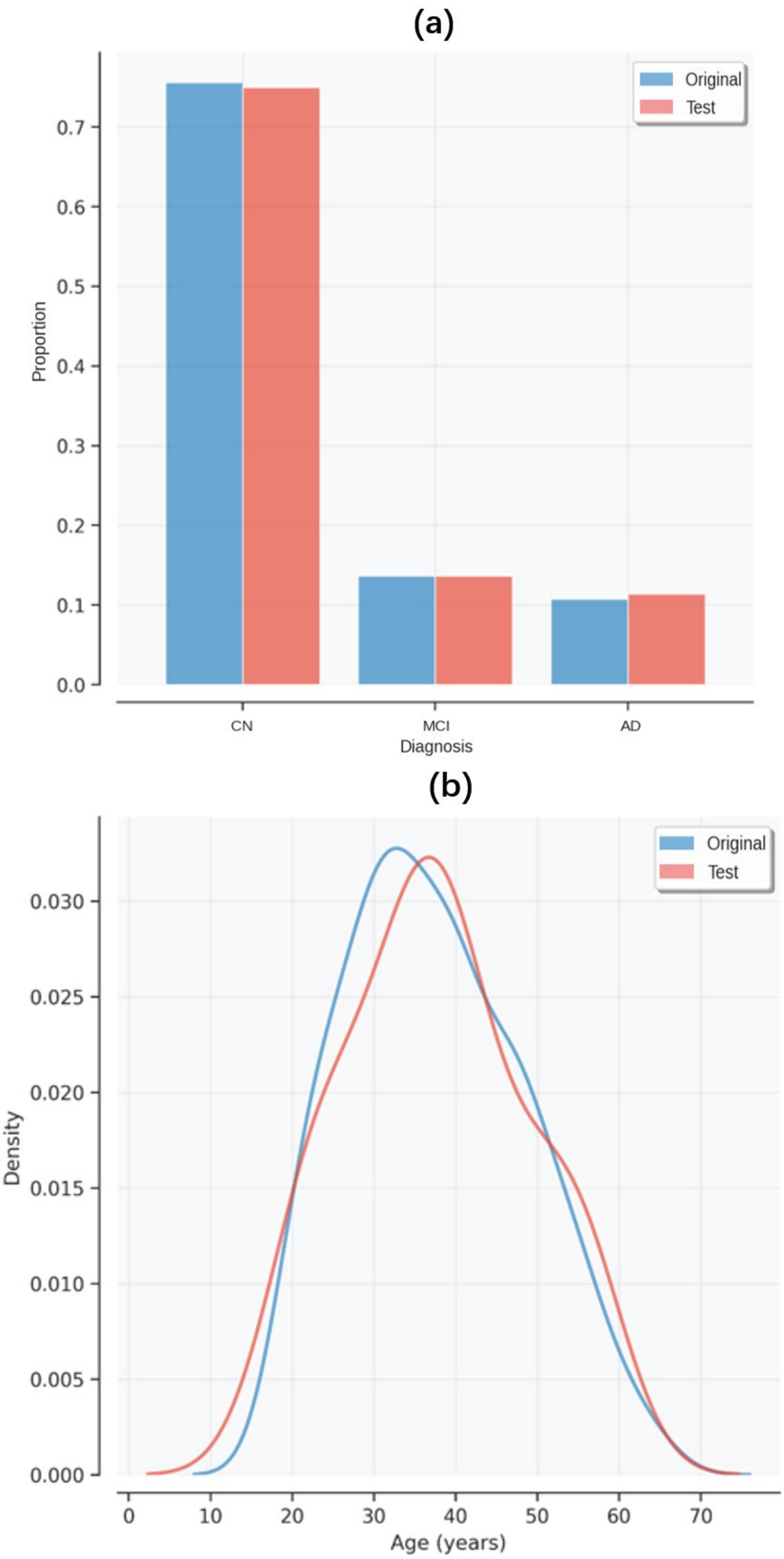

(c)

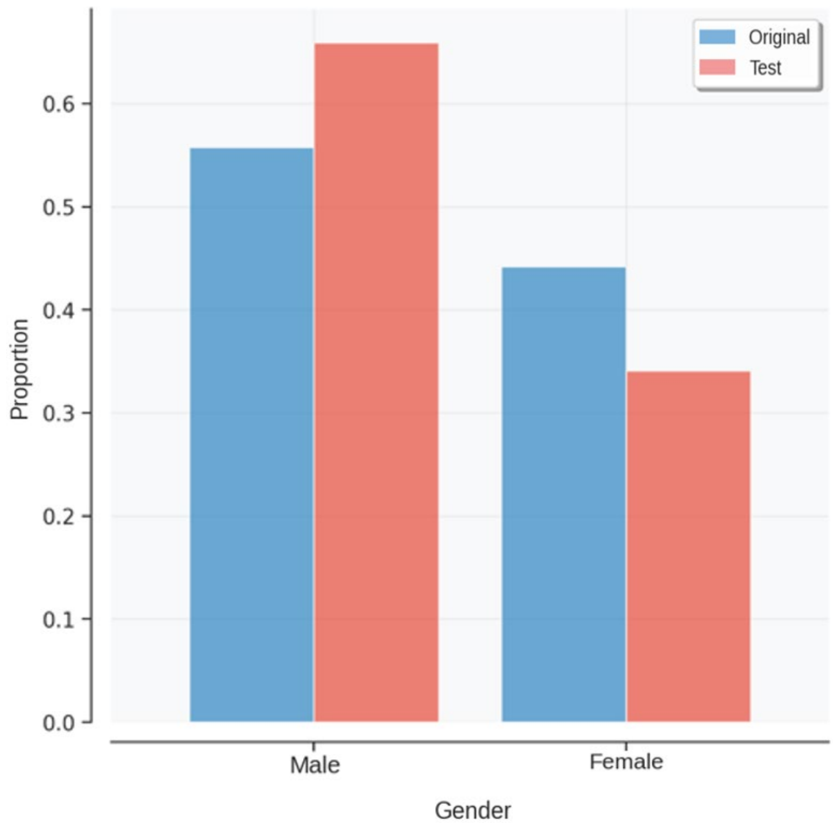


(d)

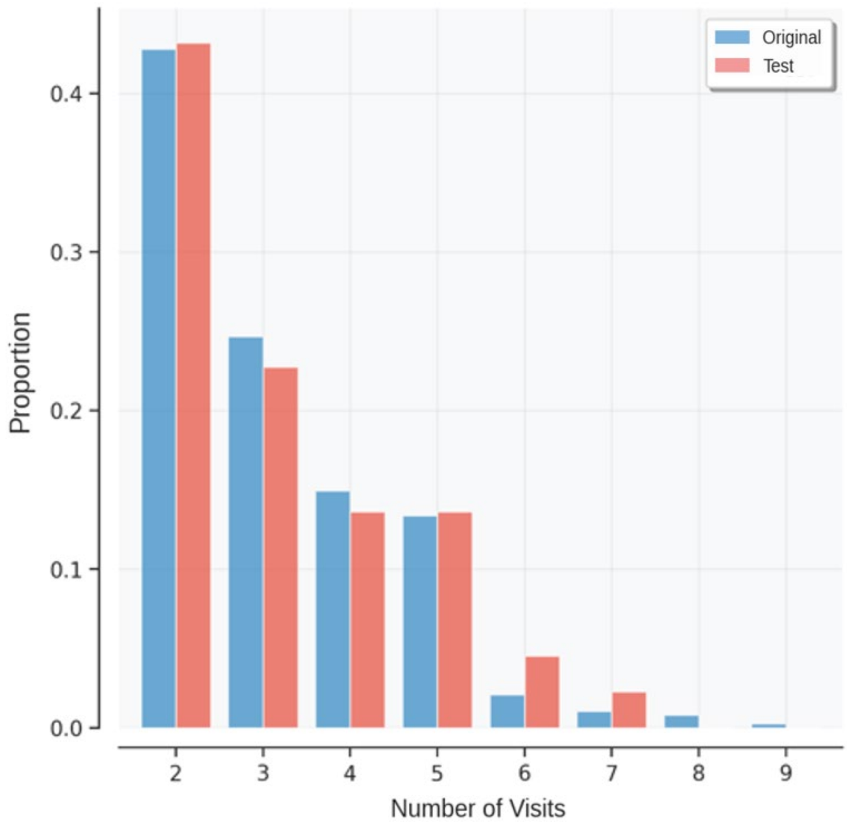


Supplementary Figure 3. Stratified data splitting strategy for DIAN dataset. (a) Diagnosis distribution in original and test sets. (b) Age distribution in original and test sets. (c) Gender distribution in original and test sets. (d) Number of clinical visits distribution in original and test sets.
